# Preoperative Serum Ferritin Level and Acute Kidney Injury after Pediatric Cardiac Surgery

**DOI:** 10.1101/2024.12.17.24319197

**Authors:** Liang Cao, Chao Xiong, Dongyun Bie, Yuan Jia, Su Yuan, Jianhui Wang

**Author notes:** Correspondence: Yuan Su, Department of Anesthesiology, Fuwai Hospital, National Center for Cardiovascular Diseases, Peking Union Medical College and Chinese Academy of Medical Sciences, Beijing, China., Address for Correspondence: No. 167, Beilishi Road, Xicheng District, Beijing 100037, China, Jianhui Wang, Department of Anesthesiology, Fuwai Hospital, National Center for Cardiovascular Diseases, Peking Union Medical College and Chinese Academy of Medical Sciences, Beijing, China. Address for Correspondence: No. 167, Beilishi Road, Xicheng District, Beijing, 100037, China. **Conflicts of Interest Statement**: Authors have nothing to disclose with regard to commercial support.

## Abstract

**Background:** Acute kidney injury (AKI) is a significant complication in pediatric patients undergoing cardiac surgery. Iron metabolism-related indicators such as ferritin may predict AKI after adult cardiac surgery. However, it remains unclear whether ferritin can be used as a predictor of AKI after pediatric cardiac surgery. This study aims to investigate the association between preoperative serum ferritin levels and the risk of AKI in pediatric population.

**Methods:** A prospective observational cohort study included 6088 pediatric patients (aged <16 years) who underwent cardiac surgery between 2022 and 2023 in Fuwai hospital. Preoperative serum ferritin levels were measured. The primary outcome was the occurrence of AKI within 7 days postoperatively, diagnosed per KDIGO criteria. Secondary outcomes included severe AKI (KDIGO stages 2 and 3), postoperative dialysis, and in-hospital mortality.

**Results:** The preoperative serum ferritin demonstrated a J-shaped association with the occurrence of AKI. Categorically, higher serum ferritin levels (>300 μ g/L, 150-300 μ g/L and 80.6-150 μ g/L) significantly increased the risk of AKI compared to lower level (40-80.6 μ g/L) (adjusted OR, 3.468; 95% CI, 1.911-6.291; adjusted OR, 3.142; 95% CI, 2.037-4.8451; and adjusted OR, 1.880; 95% CI, 1.299-2.495; respectively). The risk of stage 2 and stage 3 AKI significantly increased with higher serum ferritin categories compared to lower level (adjusted OR, 4.428; 95% CI, 1.631-12.026; adjusted OR, 3.707; 95% CI, 1.710-8.035; and adjusted OR, 2.345; 95% CI, 1.220-4.505; respectively). As a continuous variable, consistent with categorical variables, elevated serum ferritin levels (>80.6μg/L) independently predicted AKI (adjusted OR, 1.001; 95% CI, 1.000-1.002).

**Conclusions:** This study highlights the importance of preoperative serum ferritin levels in predicting AKI risk in pediatric patients undergoing cardiac surgery. Further research is warranted to elucidate the underlying mechanisms and explore the therapeutic implications of ferritin monitoring in clinical practice.

**Registration:** URL: https://www.clinicaltrials.gov; Unique identifier: NCT05489263.

**Clinical Perspective:** *What Is New?:* An important role for iron metabolism in the pathogenesis of AKI has long been appreciated. Inflammatory factors and hemodynamic and the release of labile iron, contributing to oxidation from reactive oxygen species are among the major determinants of CSA-AKI. This study highlights the importance of preoperative serum ferritin levels in predicting AKI risk in pediatric patients undergoing cardiac surgery. In this prospective observational cohort study included 3703 pediatric patients, who developed AKI had significantly higher concentrations of preoperative serum ferritin. The elevated serum ferritin levels exceeding 80.6μg/L were significantly correlated with AKI and severe AKI. This study highlights elevated preoperative serum ferritin level is an early warning indicator of CSA-AKI.

*What Are the Clinical Implications?:* Among pediatric patients undergoing cardiac surgery, serum creatine delayed diagnose of AKI. This study is the first to establish a relationship between elevated serum ferritin levels and CSA-AKI in a substantial pediatric population, thereby offering novel insights into AKI mechanisms within this demographic. Early elevation of serum ferritin may serve as an early warning sign for AKI development in pediatric patients, with implications for utilizing ferritin as a biomarker to stratify AKI risk and exploring iron chelation therapy as a preventive measure.

**Abbreviated legend for Central Picture:** 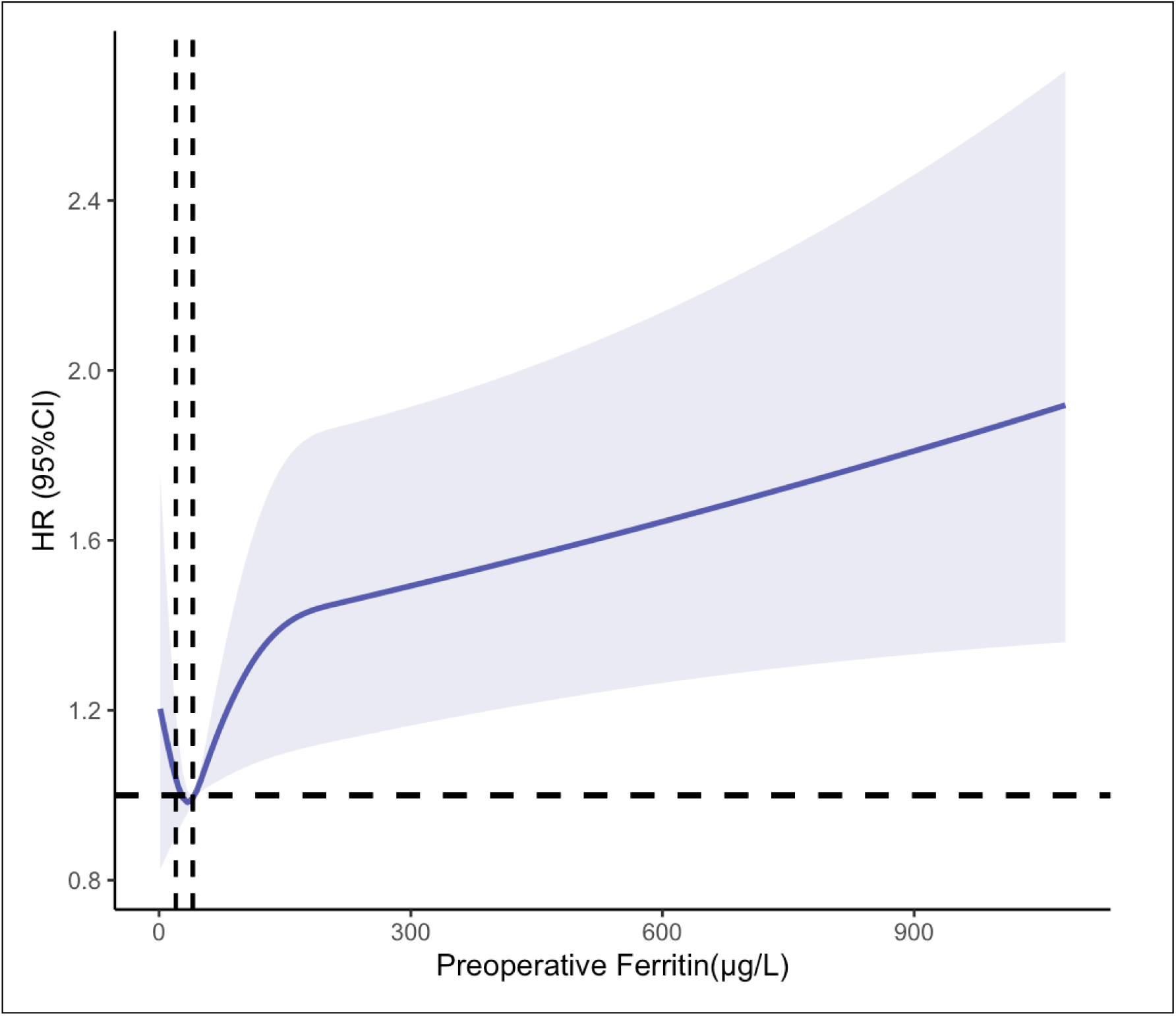

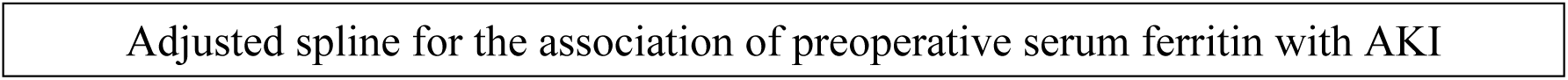

## Introduction

Acute kidney injury (AKI) is a prevalent complication following pediatric cardiac surgery, with incidence rates reported between 25% to 60%[1][2]. Cardiac surgery-associated AKI (CSA-AKI) significantly contributes to increased short– and long-term morbidity and mortality, prolonged hospital stays, and an elevated risk of developing chronic kidney disease[3][4][5][6]. While targeted preventive strategies for CSA-AKI remain underdeveloped, the timely identification of high-risk pediatric patients can facilitate proactive management.

Traditionally, AKI diagnosis relies on serum creatinine (SCr) levels and urine output measurements, which often manifest days after initial renal insult[7]. Thus, the prompt and accurate identification of pediatric patients with congenital heart disease at risk for AKI is critical for effective perioperative management.

Emerging evidence suggests that indicators of iron metabolism, such as hepcidin, catalytic iron, and serum ferritin, may correlate with AKI and other critical conditions, including acute coronary syndrome[8]. The involvement of iron metabolism in tissue injury during ischemia-reperfusion has been increasingly recognized, with several studies indicating a potential role for these biomarkers in predicting AKI following cardiac surgery[9][10][11]. However, current literature is limited by small sample sizes and inconsistent findings, particularly regarding the relationship between iron metabolism-related indicators and postoperative AKI in the pediatric population.

This prospective cohort study aims to investigate the association between levels of iron metabolism-related indicators and the incidence of AKI following pediatric cardiac surgery, as well as to assess their predictive value for AKI development.

## Methods

### Study population

We conducted a prospective, observational, single-center cohort study involving pediatric patients under 16 years of age who underwent cardiac surgery at Fuwai Hospital, Chinese Academy of Medical Sciences, Beijing, China, from January 2022 to December 2023. Exclusion criteria included: (1) age over 16 years, (2) preoperative dialysis or baseline serum creatinine >4 mg/dL, (3) unclear AKI status within the first 7 days post-surgery, (4) absence of preoperative iron metabolic data, and (5) multiple cardiac surgeries, where only the curative surgical admission was included. Institutional Review Board approval was obtained, with written consent waived, and the study is registered at https://clinicaltrials.gov/ (NCT05489263). The study followed STROBE guidelines for observational research.

## Data Collection

Preoperative data collected included age at surgery, sex, weight, left ventricular ejection fraction (LVEF), baseline serum creatinine, albumin, hemoglobin, and iron metabolism tests(serum iron, ferritin, transferrin, total iron-binding capacity [TIBC], and transferrin saturation [TAST]). Baseline laboratory values were the most recent measurements taken within 7 days prior to surgery. Intraoperative data included the Risk Adjustment for Congenital Heart Surgery 1 (RACHS-1) scoring system for surgical complexity[12], use of cardiopulmonary bypass (CPB), CPB duration, and cross-clamp time.

## Outcomes

The primary outcome was the occurrence of AKI within 7 days post-cardiac surgery, diagnosed per KDIGO criteria[13]. Secondary outcomes included severe AKI (KDIGO stages 2 and 3), postoperative dialysis, and in-hospital mortality. Given the low body weight of some patients, both peritoneal dialysis and renal replacement therapy were classified as dialysis for this analysis. Additional evaluations encompassed the duration of mechanical ventilation and lengths of ICU and hospital stays.

## Statistical Analysis

Continuous variables were presented as medians (interquartile range [IQR]) and compared between groups using the Wilcoxon rank-sum test. Categorical variables were presented as numbers (percentages) and analyzed using Pearson’s chi-square tests. For trichotomous variables, the Kruskal-Wallis test was applied for nonparametric analyses.

Restricted cubic splines were utilized to model the associations between preoperative iron metabolism indicators and postoperative AKI. The optimal threshold for serum ferritin levels predicting AKI progression was determined using receiver operating characteristic (ROC) curve analysis. The relationship between preoperative serum ferritin levels—both as continuous and categorical variables—and primary and secondary outcomes was assessed using unadjusted and adjusted logistic regression. Adjustments were made for potential confounders, including age, sex, weight, LVEF, RACHS-1 score, baseline serum creatinine, albumin, and hemoglobin. Results are presented as odds ratios (ORs) or hazard ratios (HRs) with 95% confidence intervals (CIs). Statistical significance was set at p < 0.05. Splines adjusted for covariates were used to visualize the association between iron metabolism-related indicators and AKI. Potential effect modifiers of the association between preoperative serum ferritin and AKI, including age, RACHS-1 score, and anemia, were assessed through subgroup analyses and interaction tests. Interaction terms were included in the models, with p-values <0.10 considered significant. Additionally, the association of ferritin as a categorical variable with AKI development was evaluated using multivariable logistic regression.

Sensitivity analyses included internal validation of models using 1,000 bootstrap samples and assessment of the serum ferritin-AKI relationship via Cox proportional hazards regression models. All statistical analyses were conducted using SPSS version 29 and R software version 4.3.3.

## Results

### Study cohort description

Between January 2022 and December 2023, a total of 6,088 pediatric patients undergoing cardiac surgery were enrolled in this prospective observational cohort. Following the exclusion of 2,136 patients due to missing baseline serum creatinine (n=157) or iron metabolism data (n=1,979), and 249 patients who underwent multiple surgeries (where only data from the curative surgery were included), a final cohort of 3,703 patients was analyzed (including 411 patients without cardiopulmonary bypass) (Figure 1).

**Figure 1.**
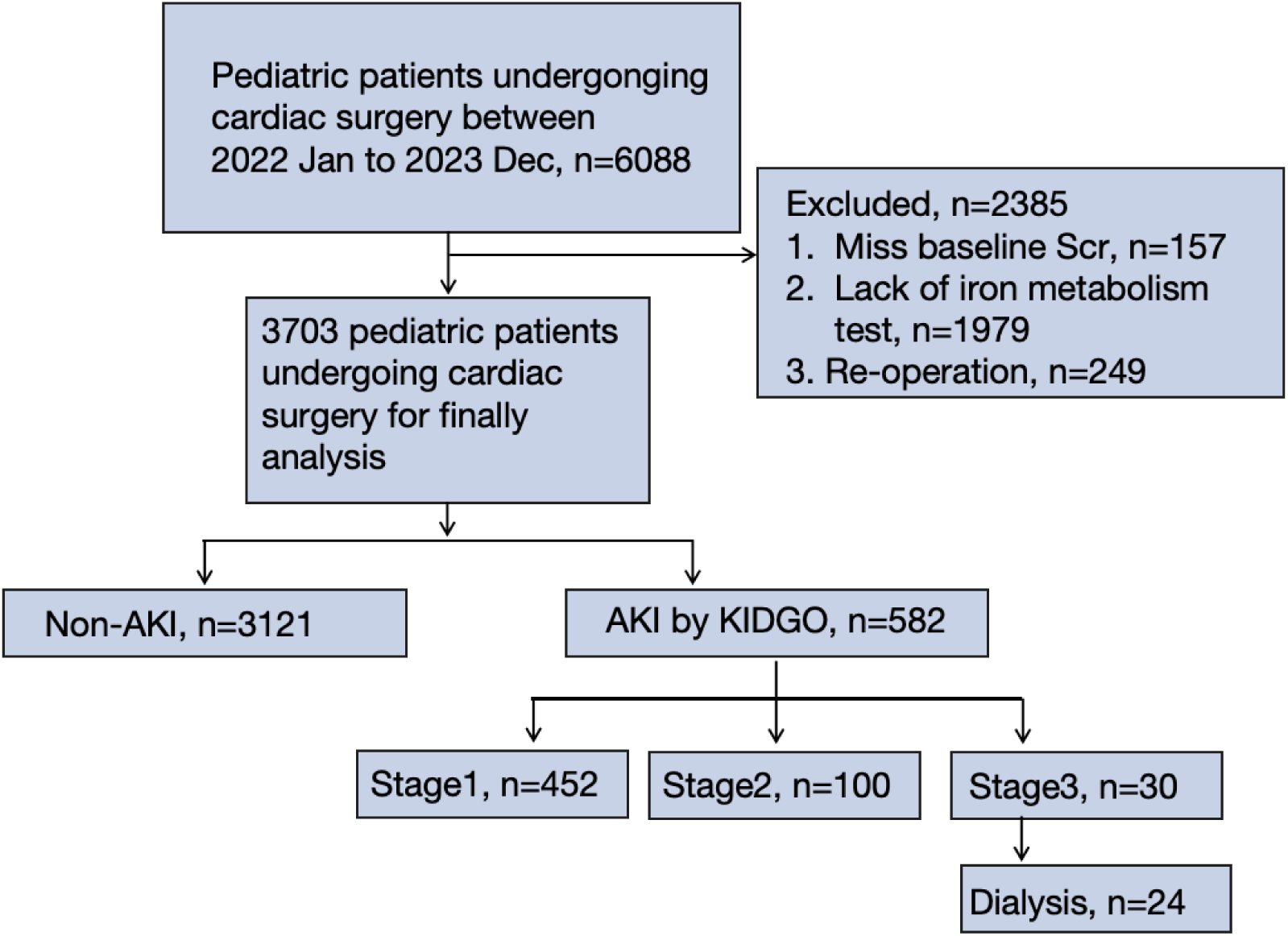
Study cohort flowchart.

The demographics, clinical characteristics, surgical information, and postoperative outcomes are summarized in Table 1. The incidence of AKI in this cohort was 15.7% (582/3703), with stage 1 AKI in 452 patients (12.2%), stage 2 in 100 (2.7%), and stage 3 in 30 (0.8%). Postoperative dialysis was required for 24 patients (0.6%) while in-hospital mortality was 0.3% (9 of 3,703), and ECMO was utilized in 0.26% (8 of 3,703). Patients who developed AKI experienced prolonged ventilation times, extended ICU and hospital stays, and higher rates of dialysis, ECMO, and mortality (Table 1).

**Table 1.**
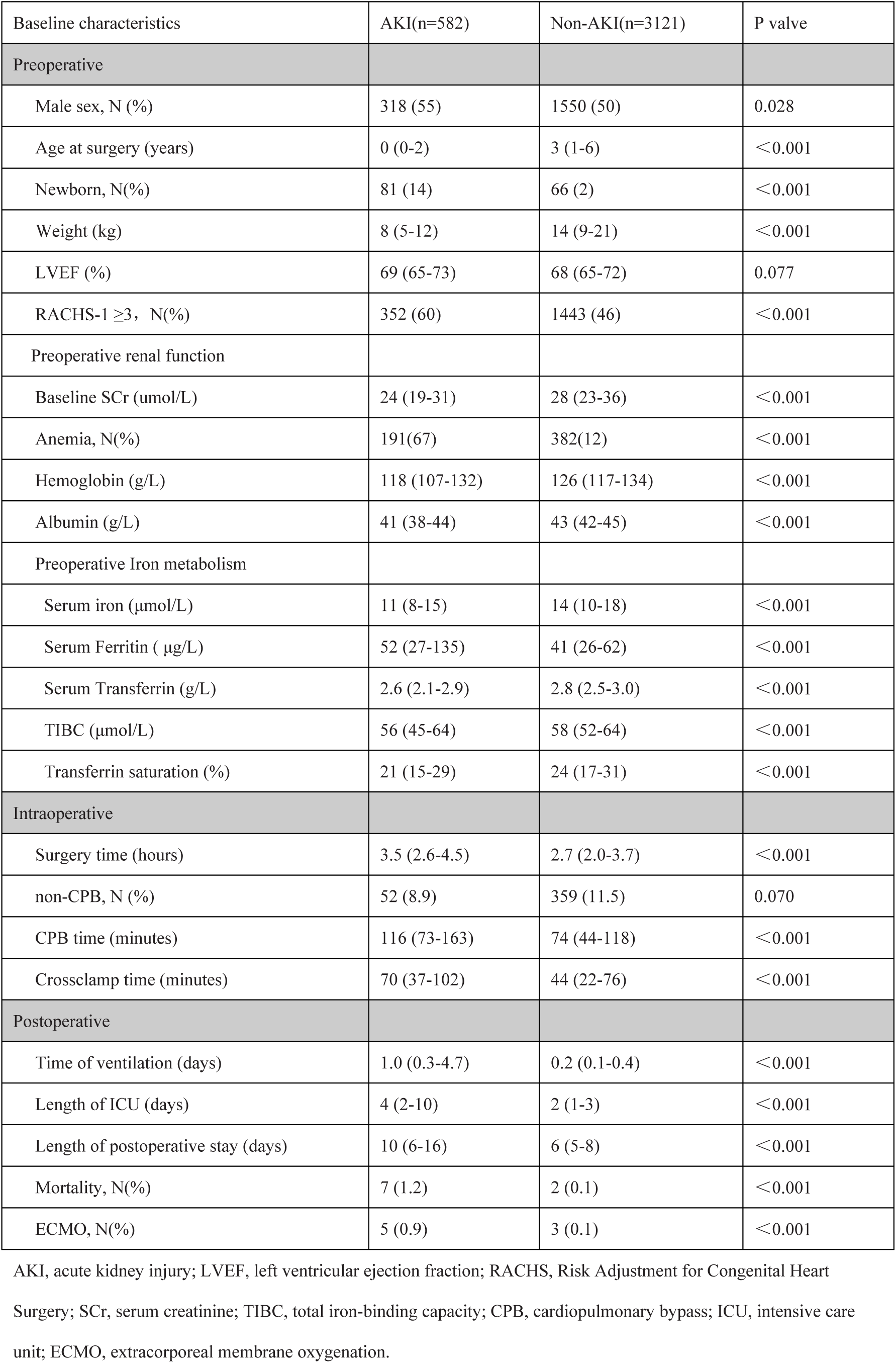
Baseline and perioperative characteristics of patients with and without acute kidney injury.

| Baseline characteristics | AKI(n=582) | Non-AKI(n=3121) | P valve |
| --- | --- | --- | --- |
| Preoperative |  |  |  |
| Male sex, N (%) | 318 (55) | 1550 (50) | 0.028 |
| Age at surgery (years) | 0 (0-2) | 3 (1-6) | <0.001 |
| Newborn, N(%) | 81 (14) | 66 (2) | <0.001 |
| Weight (kg) | 8 (5-12) | 14 (9-21) | <0.001 |
| LVEF (%) | 69 (65-73) | 68 (65-72) | 0.077 |
| RACHS-1 $\geq 3$, N(%) | 352 (60) | 1443 (46) | <0.001 |
| Preoperative renal function |  |  |  |
| Baseline SCr (umol/L) | 24 (19-31) | 28 (23-36) | <0.001 |
| Anemia, N(%) | 191(67) | 382(12) | <0.001 |
| Hemoglobin (g/L) | 118 (107-132) | 126 (117-134) | <0.001 |
| Albumin (g/L) | 41 (38-44) | 43 (42-45) | <0.001 |
| Preoperative Iron metabolism |  |  |  |
| Serum iron (μmol/L) | 11 (8-15) | 14 (10-18) | <0.001 |
| Serum Ferritin ( μg/L) | 52 (27-135) | 41 (26-62) | <0.001 |
| Serum Transferrin (g/L) | 2.6 (2.1-2.9) | 2.8 (2.5-3.0) | <0.001 |
| TIBC (μmol/L) | 56 (45-64) | 58 (52-64) | <0.001 |
| Transferrin saturation (%) | 21 (15-29) | 24 (17-31) | <0.001 |
| Intraoperative |  |  |  |
| Surgery time (hours) | 3.5 (2.6-4.5) | 2.7 (2.0-3.7) | <0.001 |
| non-CPB, N (%) | 52 (8.9) | 359 (11.5) | 0.070 |
| CPB time (minutes) | 116 (73-163) | 74 (44-118) | <0.001 |
| Crossclamp time (minutes) | 70 (37-102) | 44 (22-76) | <0.001 |
| Postoperative |  |  |  |
| Time of ventilation (days) | 1.0 (0.3-4.7) | 0.2 (0.1-0.4) | <0.001 |
| Length of ICU (days) | 4 (2-10) | 2 (1-3) | <0.001 |
| Length of postoperative stay (days) | 10 (6-16) | 6 (5-8) | <0.001 |
| Mortality, N(%) | 7 (1.2) | 2 (0.1) | <0.001 |
| ECMO, N(%) | 5 (0.9) | 3 (0.1) | <0.001 |

AKI, acute kidney injury; LVEF, left ventricular ejection fraction; RACHS, Risk Adjustment for Congenital Heart
Surgery; SCr, serum creatinine; TIBC, total iron-binding capacity; CPB, cardiopulmonary bypass; ICU, intensive care
unit; ECMO, extracorporeal membrane oxygenation.

### Associations between preoperative iron metabolism indicators and AKI

Preoperative biomarkers of iron status demonstrated significant associations with postoperative AKI. Patients who developed AKI exhibited lower levels of preoperative serum iron [11 (8-15) vs. 14 (10-18) μ mol/L, p < 0.001], serum transferrin [2.6 (2.1-2.9) vs. 2.8 (2.5-3.0) g/L, p < 0.001], total iron-binding capacity (TIBC) [56 (45-64) vs. 58 (52-64) μ mol/L, p < 0.001], and transferrin saturation (TAST) [21 (15-29) vs. 24 (17-31)%, p < 0.001] (Table 1). Notably, serum iron, transferrin, TIBC, and TAST progressively decreased with increasing AKI stage (Figure 2). Multivariable analyses utilizing restricted cubic splines revealed a relationship between these iron parameters and AKI (Figures 3).

**Figure 2.**
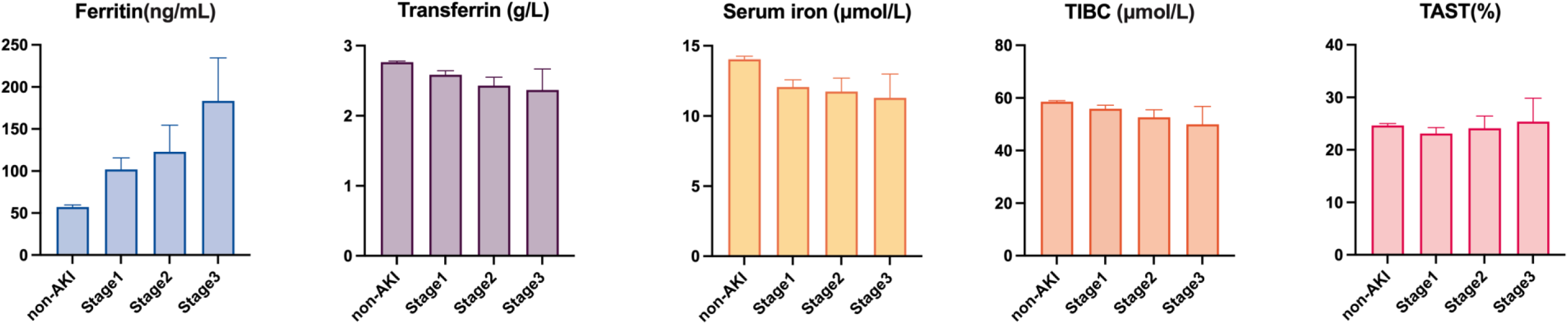
Distribution of iron parameters across different stages of AKI Stages 1 (n=452), Stages 2 (n=100), Stages 3 (n=30) and non-AKI (n=3121). Height of the bar chart shows the mean value and error bar shows the standard error.

**Figure 3.**
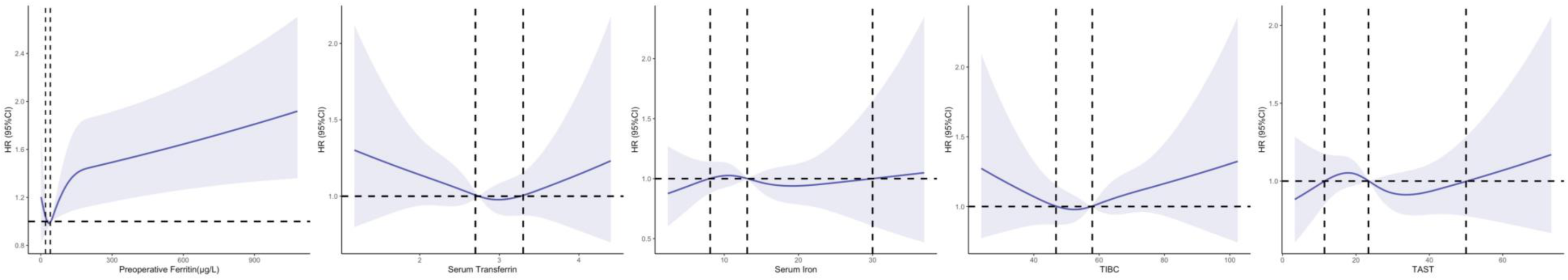
Adjusted spline for the association of preoperative iron parameters with acute kidney injury after cardiac surgery. Spline adjusted for age, sex, weight, LVEF, baseline albumin, baseline serum creatinine, baseline hemoglobin and RACHS-1 score. LVEF, left ventricular ejection fraction; RACHS, Risk Adjustment for Congenital Heart Surgery; HR, hazard rate

### Associations between preoperative serum ferritin and AKI

The median preoperative serum ferritin level was 42.1 μ g/L (25^th^-75^th^ percentiles: 26.3-67.3 μ g/L). Patients who developed AKI had significantly elevated preoperative serum ferritin levels [52 (27-135) vs. 41 (26-62) μ g/L, p < 0.001]. Multivariable analyses revealed a J-shaped relationship between preoperative serum ferritin and AKI (Figure 2), indicating that the odds of postoperative AKI increased monotonically for ferritin levels above 40 μ g/L. The area under the ROC curve for preoperative serum ferritin was 0.596 (95% CI: 0.567 to 0.625), with an optimal cutoff identified at 80.6 μ g/L (Supplement Fig 2 and Supplement Table 2). Patients were categorized based on ferritin levels: >80.6 μ g/L, 150-300 μ g/L, and >300 μ g/L, with a reference group of 40-80.6 μ g/L. Those with ferritin <40 μ g/L were compared with a reference of 20-40 μ g/L.

Demographics and clinical characteristics stratified by serum ferritin levels are presented in Supplement Table 1. Higher ferritin levels were associated with younger age, lower weight, increased surgical complexity, and elevated baseline serum creatinine, along with reduced hemoglobin and albumin levels. A positive correlation between elevated ferritin levels and increased AKI incidence and dialysis requirement was observed.

As a continuous variable, higher serum ferritin was linked to an increased risk of developing AKI (unadjusted OR, 1.003; 95% CI, 1.001-1.004; p < 0.001; Table 2). This association persisted in a multivariable logistic regression model (adjusted OR, 1.001; 95% CI, 1.000-1.002; p < 0.001). As a categorical variable, a significant trend of increasing AKI incidence was noted with rising ferritin levels (p < 0.001). After adjustments, patients with ferritin levels >300 μ g/L had a notably higher risk of AKI compared to those with 40-80.6 μ g/L (adjusted OR, 3.468; 95% CI, 1.911-6.291; p < 0.001). Increased risks were also identified for the 150-300 μ g/L and 80.6-150 μ g/L categories (adjusted ORs: 3.142 and 1.880, respectively; p < 0.001). (Table 2).

**Table 2.**
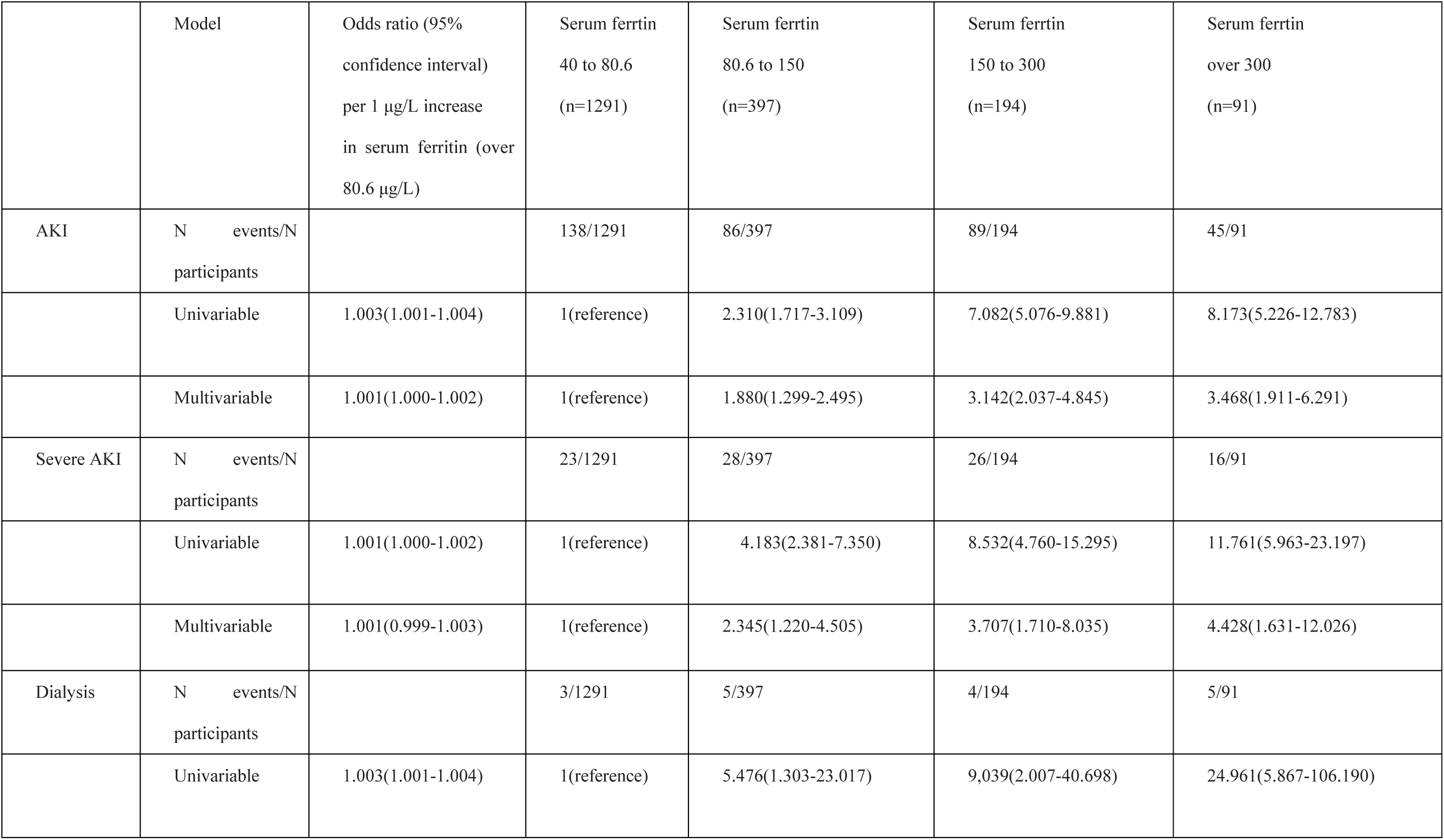
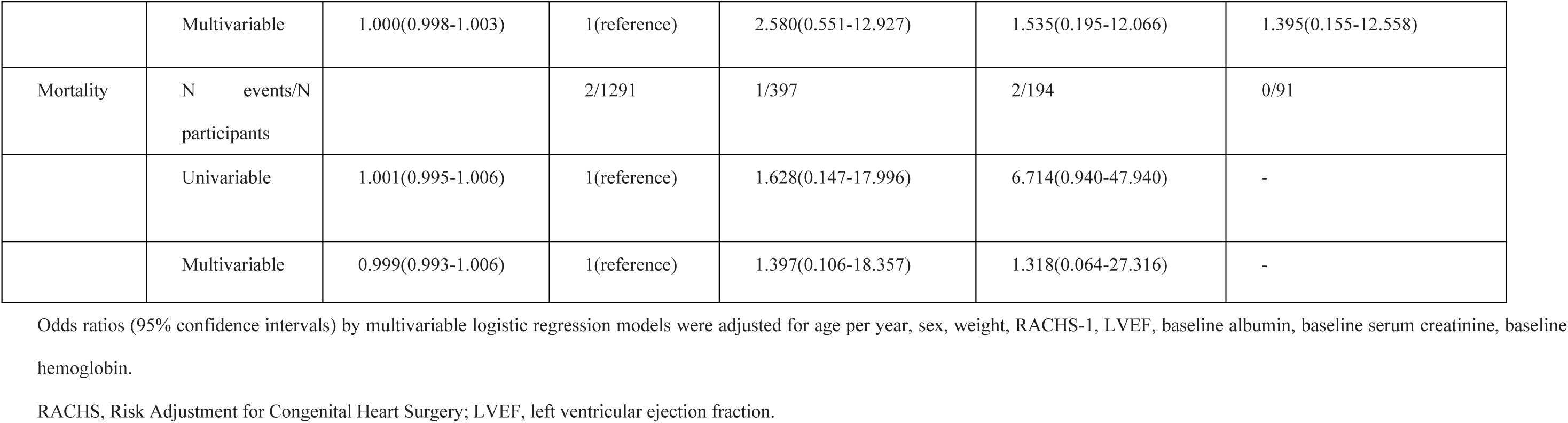
Association of serum ferritin (over 80.6 μg/L) with outcomes after cardiac surgery.

### Associations between preoperative serum ferritin and secondary outcomes

Multivariable analyses revealed a significantly higher risk of severe AKI (KDIGO stages 2 and 3) among patients with ferritin >300 μg/L, 150-300 μg/L, and 80.6-150 μg/L (adjusted ORs: 4.428, 3.707, and 2.345, respectively; p < 0.003 for all) (Table 2). A significant association was noted for the requirement of dialysis (OR:24.961, 9.039, and 5.476, respectively; p < 0.005 for all). However, this significance was lost after adjustment (adjust OR 1.395; 95% CI: 0.155-12.558). Preoperative serum ferritin levels did not significantly correlate with in-hospital mortality in multivariable analysis. For patients with ferritin <20 μg/L, no significant correlations with AKI, severe AKI, dialysis, or mortality were observed (Supplement Table 3).

### Subgroup and sensitivity analyses

Subgroup analysis based on anemia status revealed that the incidence of preoperative anemia was 67% in the AKI group compared to 12% in the non-AKI group (p< 0.001). Among pediatric patients without preoperative anemia, a significant increase in AKI risk was noted with rising ferritin levels (Table 3). An interaction between preoperative ferritin and anemia was observed (p for interaction 0.045), indicating variability across hemoglobin levels. In patients undergoing complex congenital cardiac surgeries (RACHS-1 score ≥ 3), preoperative ferritin concentrations significantly correlated with AKI occurrence (Table 3). Sensitivity analyses confirmed the robustness of these findings (Supplement Tables 4 and 5).

**Table 3.** Subgroup and interaction analyses for age, RACHS-1 score, and anemia. Subgroup analyses of the associations between preoperative serum ferritin levels and acute kidney injury in multivariable logistic regression models.

| Subgroup | Number of patients | Serum ferritin 40 to 80.6 (n=1291) | Serum ferritin 80.6 to 150 (n=397) | Serum ferritin 150 to 300 (n=194) | Serum ferritin over 300 (n=91) | p for interaction |
| --- | --- | --- | --- | --- | --- | --- |
| Age |  |  |  |  |  | 0.153 |
| newborn | 134 | 1(reference) | 1.685(0.295-9.633) | 2.439(0.469-12.679) | 3.004(0.507-17.781) |  |
| 1m to 1y | 257 | 1(reference) | 1.085(0.643-1.832) | 1.332(0.739-2.403) | 1.436(0.639-3.229) |  |
| > 1 year | 291 | 1(reference) | 1.797(1.122-2.880) | 2.080(0.747-5.791) | - |  |
| RACHS-1 score |  |  |  |  |  | 0.929 |
| <3 | 274 | 1(reference) | 1.535(0.912-2.582) | 1.815(0.884-3.726) | 2.953(1.708-8.088) |  |
| ≥3 | 408 | 1(reference) | 1.890(1.238-2.884) | 3.247(1.927-5.344) | 3.320(1.738-6.344) |  |
| Anemia |  |  |  |  |  | 0.045 |
| yes | 226 | 1(reference) | 2.317(1.064-5.045) | 1.575(0.727-3.411) | 2.650(1.018-6.898) |  |
| no | 456 | 1(reference) | 1.509(1.031-2.209) | 2.772(1.634-4.704) | 2.229(1.002-4.959) |  |

Odds ratios (95% confidence intervals) by multivariable logistic regression models were adjusted for age per year, sex, weight, RACHS-1, LVEF, baseline albumin, baseline serum creatinine, baseline hemoglobin.
RACHS, Risk Adjustment for Congenital Heart Surgery; LVEF, left ventricular ejection fraction.

## Discussions

In this prospective cohort study of 3703 pediatric patients undergoing cardiac surgery, we identified a significant association between preoperative serum ferritin levels and pediatric CSA-AKI. Notably, patients with elevated preoperative ferritin levels were at increased risk of developing postoperative AKI, particularly when ferritin exceeded 80.6 μg/L. This stepwise increase in risk remained robust after adjusting for multiple confounders, indicating the potential predictive value of ferritin levels for AKI in this population. Additionally, elevated serum ferritin levels correlated with a significantly higher likelihood of severe AKI (KDIGO stages 2 and 3).

To our knowledge, this study is the first to establish a relationship between elevated serum ferritin and CSA-AKI in pediatric patients. Our findings extend previous research on adults, which demonstrated differing associations between ferritin and kidney injury. In adult studies, higher preoperative ferritin levels have been inversely associated with AKI risk, as seen in a study involving 301 patients undergoing cardiac surgery[8]. A meta-analysis including 451 patients indicated that those developing AKI after cardiac surgery had lower baseline serum ferritin levels[10].

Furthermore, another study of 250 adult patients who underwent cardiac surgery showed that while preoperative ferritin was not significantly associated with renal replacement therapy or mortality, postoperative ferritin levels were significantly elevated among patients who did experience these outcomes on postoperative day 1 (OR: 2.09; 95% CI, 1.33-3.31; P<0.01)[9]. These discrepancies highlight potential age-related or physiological differences between pediatric and adult patients in the context of ferritin’s role in AKI.

Iron overload due to factors such as blood transfusions and hemolysis can elevate the risk of kidney disease. Increasing serum levels of catalytic iron, ferritin and hepcidin are associated with progressive worsening of long-term outcomes in patients with mild-to-moderate chronic kidney disease[14]. The deleterious effects of labile iron include the generation of free radicals and oxidative injury, particularly affecting the cardiovascular and renal systems[15]. Mechanisms such as disruption of cell-matrix adhesion and interference with the proliferation of renal tubular epithelial cells can cause kidney injury[16]. Ferritin, an iron storage protein, can mop up excess iron; on another hand, it may serve as an additional source of catalytic iron by releasing it in response to inflammation-induced superoxide production or ischemia-reperfusion injury, contributing to AKI risk. Catalytic iron has been implicated in a higher risk of AKI and death in the setting of cardiac surgery and in critically ill subjects[10][17]. In patients with established AKI, higher catalytic iron was found to be associated with about a four-fold increased risk of death[18].

In pediatric patients, elevated serum ferritin levels prior to surgery suggest two potential mechanisms. First, as an acute-phase reactant, ferritin can rise in response to inflammation, particularly evident in pediatrics with severe conditions undergoing complex congenital cardiac surgery. Our subgroup analysis indicated a more pronounced effect of ferritin on AKI risk among patients with higher RACHS-1 scores. Preoperative inflammation may exacerbate renal hypoxia, especially further progress after prolonged surgeries involving cardiopulmonary bypass. Second, during the stress response to surgery, ferritin as an iron storage protein may release catalytic iron, leading to oxidative damage via the Fenton reaction, especially in the kidney.

The association of serum ferritin with AKI implies that iron regulatory pathways may be activated in response to ischemia-reperfusion injury during cardiopulmonary bypass, potentially exacerbating the impact of catalytic iron release[19][20]. Elevated preoperative ferritin levels appear to increase AKI risk secondary to the effects of cardiopulmonary bypass, as the protective mechanisms of the heme oxygenase system may be saturated, leaving no reserve capacity to counteract the injuries sustained during such procedures[21][22].

Several studies have demonstrated the pivotal role of ferroptosis in the development of AKI following cardiac surgery[23][24][25][26]. Post-surgical release of a substantial amount of catalytic iron has been implicated as a potential trigger for AKI through the induction of ferroptosis[27]. Catalytic iron can participate in the Fenton reaction, generating reactive oxygen species (ROS) that contribute to oxidative stress. This oxidative stress is a key mechanism in the induction of ferroptosis, a regulated form of cell death characterized by lipid peroxidation. Conversely, excessive iron released from other sources like free hemoglobin may stimulate elevated levels of ferritin as a protective chelating mechanism[28]. Dysregulated serum iron levels can upregulate hepcidin expression, thereby reducing serum iron concentrations[29]. Therefore, ferritin or hepcidin could potentially be a therapeutic target for AKI after cardiac surgery.

We identified a significant association between elevated serum ferritin levels and AKI in pediatric patients without preoperative anemia, despite anemia being a recognized risk factor for AKI[30][31]. Although ferritin decreases earlier than hemoglobin, we did not observe a significant influence of decreased preoperative serum ferritin on postoperative AKI.

Although we cannot establish causality from observational study, early elevation of serum ferritin may serve as an early warning sign for AKI development in pediatric patients, with implications for utilizing ferritin as a biomarker to stratify AKI risk and exploring iron chelation therapy as a preventive measure.

## Study Strengths and Limitations

Our study possesses several notable strengths. This study is the first to establish a relationship between elevated serum ferritin levels and CSA-AKI in a substantial pediatric population, thereby offering novel insights into AKI mechanisms within this demographic. The prospective observational design enabled a comprehensive analysis characterized by meticulously collected data with minimal missing values, well-defined outcome measures, and continually assessment of AKI within seven days postoperatively.

However, certain limitations must be acknowledged. Firstly, the findings stem from a single-center cohort, which may restrict the generalizability of the results; validation through multicenter studies is warranted. Secondly, the observational nature of the study limits our ability to infer causality between elevated ferritin levels and AKI progression. While the data suggest that changes in serum ferritin may reflect distinct iron homeostatic mechanisms that could confer renal protection, the specific role of ferritin as either a source or mitigator of elevated catalytic iron levels remains unresolved, necessitating further research to establish causation. Additionally, the relatively low incidence of dialysis and mortality within the cohort may limit the applicability of exploring these relationships. Nevertheless, preoperative elevated ferritin levels demonstrated superior predictive capability compared to serum creatinine for postoperative AKI and proved to be more readily accessible than previously reported intraoperative urine and plasma biomarkers for clinical diagnosis[32]. Importantly, further investigation is required to assess whether serial monitoring of ferritin levels might provide greater predictive value for outcomes than relying on baseline levels alone.

## Conclusions

In conclusion, this study establishes a significant association between elevated preoperative serum ferritin levels and the risk of AKI in pediatric patients undergoing cardiac surgery. Our findings suggest that serum ferritin may serve as a valuable biomarker for identifying patients at increased risk of AKI, offering potential for early intervention strategies. The observed relationship underscores the need for further investigation into the mechanistic role of ferritin in renal injury, particularly concerning its interactions with catalytic iron and iron homeostasis. Given the implications for clinical practice, future multicenter studies are essential to validate these findings and explore the therapeutic potential of targeting ferritin in mitigating AKI risk in this vulnerable population.

## Conflict of Interest Statement

This study was supported by the Chinese Academy of Medical Sciences Central Public Welfare Scientific Research Institute Basal Research Expenses-Clinical and Translational Medicine Research Fund (2021-I2M-C&T-B-036).

**Supplementary Fig 1.**
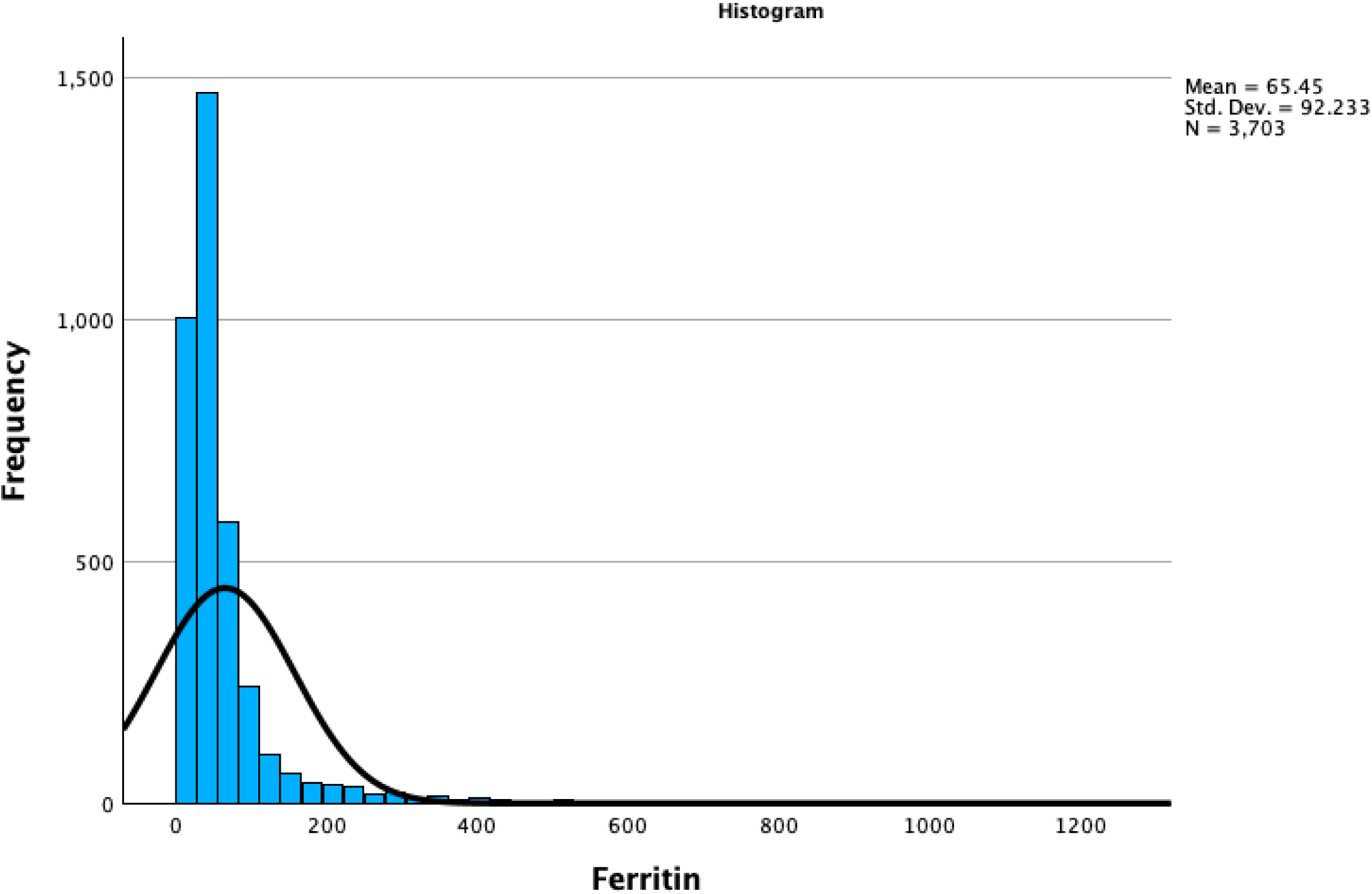
Distribution of peroperative serum ferritim.

**Supplement Fig 2.**
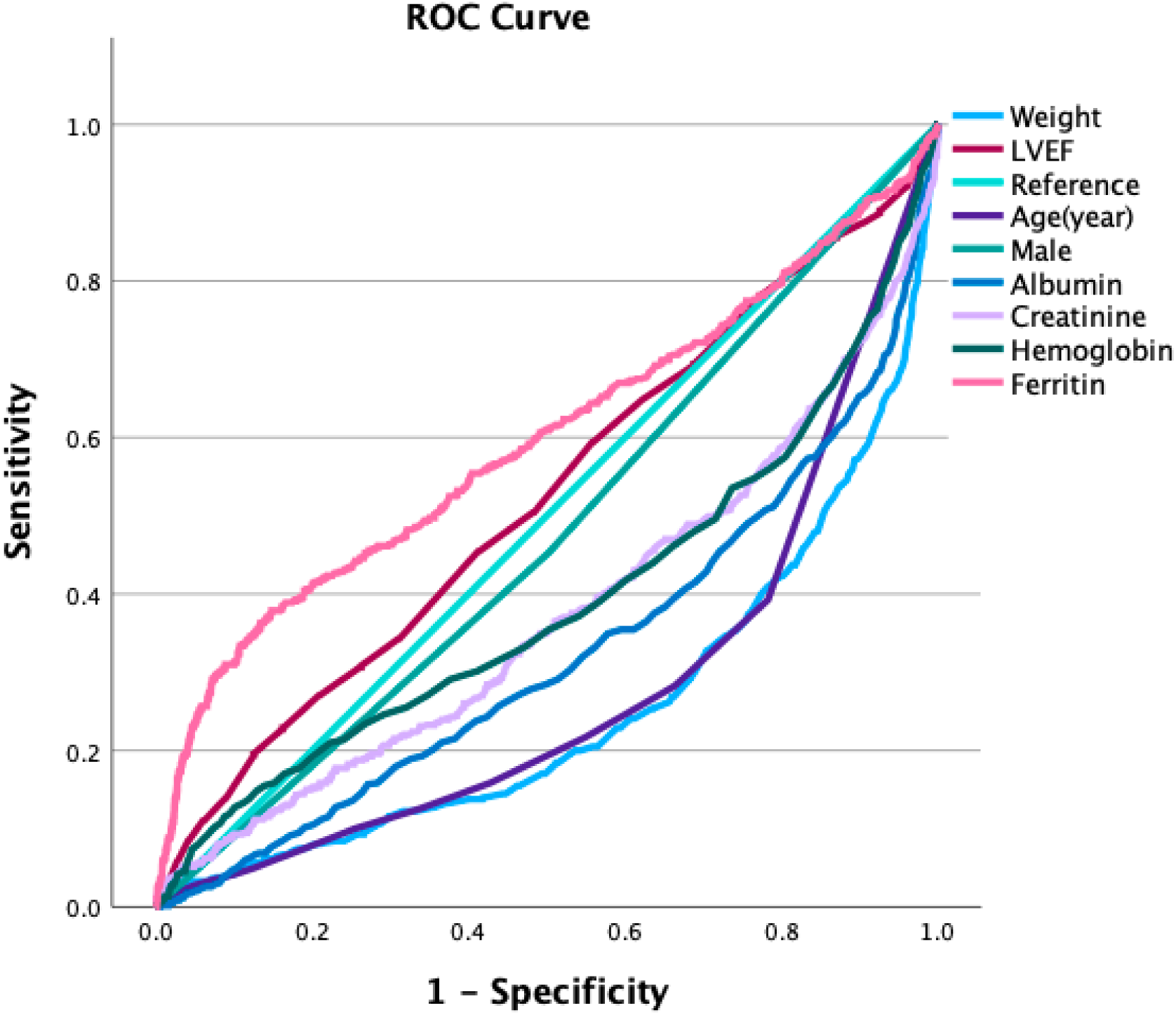
ROC curve of preoperative characteristics with development of acute kidney injury. ROC, receiver operating characteristics curve

**Supplement Table 1.** Baseline and clinical characteristics according to preoperative serum ferrtin levels.

| Characteristics | Serum ferritin<br>less than 20<br>(n=565) | Serum ferritin<br>20 to 40<br>(n=1165) | Serum ferritin 40<br>to 80.6<br>(n=1291) | Serum ferritin<br>80.6 to 150<br>(n=397) | Serum ferritin<br>150 to 300<br>(n=194) | Serum ferritin<br>over 300<br>(n=91) |
| --- | --- | --- | --- | --- | --- | --- |
| Preoperative |  |  |  |  |  |  |
| Male sex, N (%) | 316(56) | 610(52) | 589(46) | 199 (50) | 111 (57) | 43 (47) |
| Age at surgery (years) | 1(0-4) | 3(1-5) | 2(1-5) | 3 (1-5) | 0 (0-0) | 0 (0-0) |
| Newborn, N(%) | 1(0.2) | 4(0.3) | 8(0.6) | 27(7) | 65(33) | 42(46) |
| Weight (kg) | 10(8-17) | 15(10-20) | 16(11-23) | 14 (9-20) | 5 (4-7) | 4 (3-5) |
| LVEF (%) | 69(65-72) | 68(65-72) | 68(65-72) | 69 (65-72) | 68 (64-72) | 69 (60-75) |
| RACHS-1≥3, N(%) | 249(44) | 511(44) | 627(49) | 212 (53) | 131 (68) | 65 (71) |
| Preoperative renal function |  |  |  |  |  |  |
| Baseline Cr (umol/L) | 25(20-33) | 27(22-33) | 28(23-36) | 27 (22-34) | 27 (20-36) | 30 (22-42) |
| Hemoglobin (g/L) | 119(107-129) | 125(118-133) | 127(119-135) | 125 (117-133) | 118 (105-136) | 114 (103-132) |
| Anemia, N(%) | 141(25) | 109(9) | 97(8) | 76(19) | 92(47) | 58(64) |
| Albumin (g/L) | 43(41-45) | 43(42-45) | 44(42-46) | 43 (41-45) | 38 (33-42) | 37 (32-40) |
| Preoperative Iron metabolism |  |  |  |  |  |  |
| Serum iron (μmol/L) | 9(6-14) | 14(10-17) | 15(11-18) | 13 (9-18) | 11 (8-14) | 11 (8-15) |
| Serum Ferritin ( μg/L) | 13(8-17) | 31(25-35) | 53(46-64) | 36 (23-51) | 197 (168-235) | 409 (349-514) |
| Serum Transferrin (g/L) | 3.1(2.8-3.4) | 2.8(2.6-3.0) | 2.7(2.5-2.9) | 2.6 (2.3-2.9) | 2.1 (1.7-2.5) | 1.8 (1.5-2.1) |
| TIBC (μmol/L) | 67(60-75) | 59(54-64) | 57(52-62) | 54 (48-60) | 44 (35-53) | 38 (32-44) |
| Transferrin saturation (%) | 14(8-22) | 23(17-29) | 26(20-32) | 26 (18-34) | 26 (20-37) | 32 (22-42) |
| Intraoperative |  |  |  |  |  |  |
| Surgery time (hours) | 2.8(2.0-3.8) | 2.7(2.0-3.6) | 2.8(2.0-3.7) | 2.7 (2.0-3.7) | 3.4 (2.5-4.2) | 3.4 (2.2-4.3) |
| non-CPB, N (%) | 62(11) | 105(9) | 150(11.6) | 37 (9.3%) | 32 (16.5%) | 25 (27.5%) |
| CPB time (minutes) | 78(45-128) | 73(45-116) | 78(45-120) | 75 (45-119) | 111(60-144) | 97 (0-146) |
| Crossclamp time (minutes) | 50(25-85) | 44(23-75) | 45(21-78) | 45 (22-77) | 67 (34-91) | 61 (0-84) |
| Postoperative |  |  |  |  |  |  |
| Time of ventilation (days) | 0.2(0.1-0.8) | 0.2(0.1-0.3) | 0.2(0.1-0.4) | 0.3 (0.2-1.0) | 2.8 (0.7-6.0) | 4.5 (2.0-7.9) |
| Length of ICU (days) | 2(1-4) | 2(1-3) | 2(1-3) | 3 (1-5) | 10 (7-20) | 20 (7-20) |
| Length of postoperative stay (days) | 6(5-10) | 6(5-8) | 6(5-8) | 7 (5-11) | 13 (9-20) | 13 (9-19) |
| Mortality, N(%) | 3(0.5) | 1(0.1) | 2(0.2) | 1 (0.3%) | 2 (1.0%) | 0 |
| ECMO, N(%) | 1(0.2) | 1(0.1) | 2(0.2) | 1 (0.3%) | 2 (1.0%) | 1 (1.1%) |
| Dialysis, N(%) | 6(1.1) | 1(0.1) | 3(0.2) | 5 (1.3%) | 4 (2.1%) | 5 (5.5%) |
| AKI, N(%) | 91(16) | 133(11) | 138(11) | 86 (22) | 89 (46) | 45 (50) |
| AKI stage2-3, N(%) | 12(2.1) | 25(2.1) | 23(1.8) | 28 (7) | 26 (13) | 16 (18) |

AKI, acute kidney injury; LVEF, left ventricular ejection fraction; RACHS, Risk Adjustment for Congenital Heart Surgery; SCr, serum creatinine; TIBC, total iron-binding capacity; CPB, cardiopulmonary bypass; ICU, intensive care unit; ECMO, extracorporeal membrane oxygenation.

**Supplement Table 2.** Association between preoperative characteistics and postoperative AKI.

| Characteristics | OR for AKI prediction | 95% CI | AUC | 95% CI | cut-off value |
| --- | --- | --- | --- | --- | --- |
| Serum Ferritin (µg/L) | 1.005 | 1.004-1.006 | 0.596 | 0.567-0.625 | 80.6 |
| Newborn | 7.69 | 5.49-10.78 | 0.563 | 0.533-0.592 |  |
| LVEF | 1.00 | 0.99-1.01 | 0.523 | 0.496-0.550 |  |
| Male | 1.22 | 1.02-1.46 | 0.477 | 0.450-0.505 |  |
| Baseline Hemoglobin (g/L) | 0.98 | 0.97-0.99 | 0.391 | 0.362-0.419 |  |
| Baseline SCr (umol/L) | 0.96 | 0.95-0.97 | 0.374 | 0.347-0.402 |  |
| Baseline Albumin (g/L) | 0.84 | 0.82-0.86 | 0.325 | 0.299-0.351 |  |
| Age (years) | 0.77 | 0.74-0.81 | 0.278 | 0.225-0.301 |  |
| Weight (kg) | 0.91 | 0.89-0.92 | 0.252 | 0.228-0.275 |  |

OR, Odds ratios; CI, confidence intervals; LVEF, left ventricular ejection fraction; SCr, serum creatinine.

**Supplement Table 3.** Association of serum ferritin ( less than 20 μg/L) with acute kidney injury after cardiac surgery.

|  | Model | Odds ratio (95%<br>confidence interval)<br>per 1 µg/L decrease<br>in serum ferritin (less than 20 µg/L) | Serum ferritin<br>20 to 40<br>(n=1165) | Serum ferritin<br>less than 20<br>(n=565) |
| --- | --- | --- | --- | --- |
| AKI | N events/N participants |  | 133/1165 | 91/565 |
|  | Univariable | 1.008 (0.972-1.045) | 1(reference) | 1.490(1.117-0.987) |
|  | Multivariable | 1.014 (0.975-1.055) | 1(reference) | 1.229(0.908-1.662) |
| Severe AKI | N events/N participants |  | 25/1165 | 12/565 |
|  | Univariable | 0.999(0.993-1.070) | 1(reference) | 0.990(0.493-1.984) |
|  | Multivariable | 1.057(0.963-1.160) | 1(reference) | 0.802(0.387-1.662) |
| Dialysis | N events/N participants |  | 1/1165 | 6/565 |
|  | Univariable | 0.888(0.743-1.062) | 1(reference) | 12.492(1.501-104.025) |
|  | Multivariable | - | 1(reference) | 7.507(0.783-71.971) |
| Mortality | N events/N participants |  | 1/1165 | 3/565 |
|  | Univariable | 1.029(0.786-1.348) | 1(reference) | 6.214(0.645-59.868) |
|  | Multivariable | 1.133(0.780-1.643) | 1(reference) | 8.972(0.719-112.001) |

**Supplement Table 4.** Association of serum ferritin (over 80.6 μg/L) with acute kidney injury after cardiac surgery using Cox proportional hazard regression model.

|  | Model | Odds ratio (95% confidence interval)<br>per 1 µg/L increase<br>in serum ferritin (over<br>80.6 µg/L) | Serum ferritin<br>40 to 80.6<br>(n=1291) | Serum ferritin<br>80.6 to 150<br>(n=397) | Serum ferritin<br>150 to 300<br>(n=194) | Serum ferritin<br>over 300<br>(n=91) |
| --- | --- | --- | --- | --- | --- | --- |
| AKI | N events/N<br>participants |  | 138/1291 | 86/397 | 89/194 | 45/91 |
|  | Univariable | 1.000(1.000-1.001) | 1(reference) | 1.431(1.089-1.881) | 1.889(1.431-2.495) | 1.798(1.267-2.551) |
|  | Multivariable | 1.000(1.000-1.001) | 1(reference) | 1.292(0.971-1.718) | 1.611(1.145-2.267) | 1.604(1.029-2.500) |

Odds ratios (95% confidence intervals) by multivariable logistic regression models were adjusted for age per year, sex, weight, RACHS-1, LVEF, baseline albumin, baseline serum creatinine, baseline
hemoglobin.
RACHS, Risk Adjustment for Congenital Heart Surgery; LVEF, left ventricular ejection fraction.

**Supplement Table 5.**
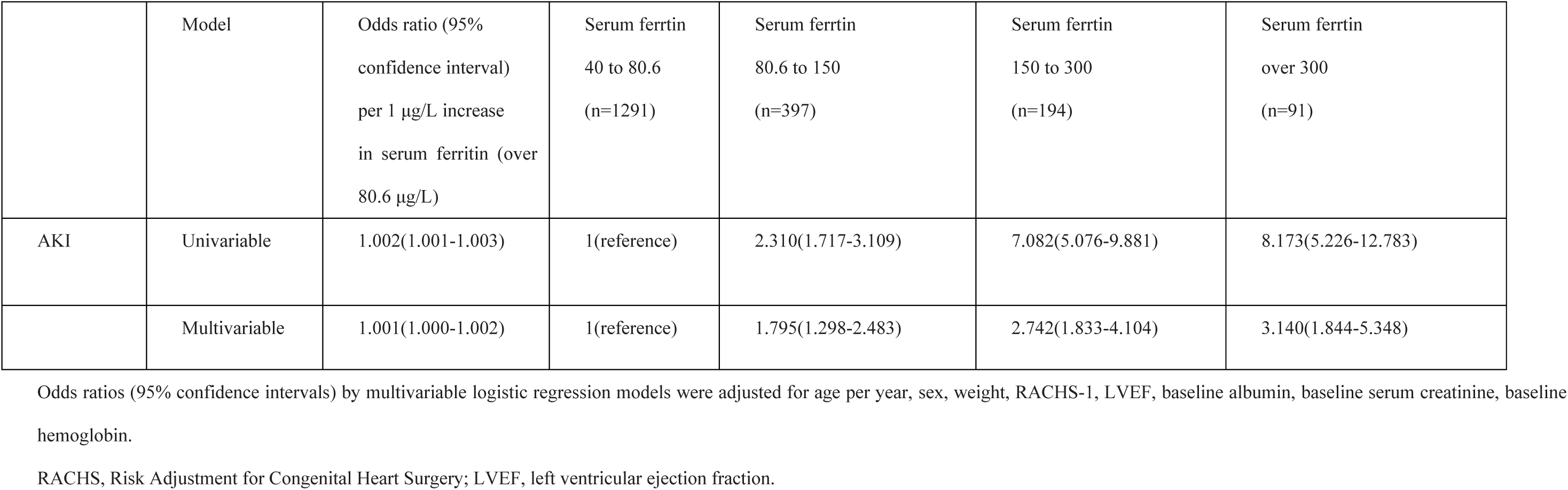
Association of serum ferritin (over 80.6 μg/L) with acute kidney injury after cardiac surgery by mutivariable logisitic regression model. This model was internally validated using 1000 bootstrap samples

|  | Model | Odds ratio (95% confidence interval) per 1 µg/L increase in serum ferritin (over 80.6 µg/L) | Serum ferritin 40 to 80.6 (n=1291) | Serum ferritin 80.6 to 150 (n=397) | Serum ferritin 150 to 300 (n=194) | Serum ferritin over 300 (n=91) |
| --- | --- | --- | --- | --- | --- | --- |
| AKI | Univariable | 1.002(1.001-1.003) | 1(reference) | 2.310(1.717-3.109) | 7.082(5.076-9.881) | 8.173(5.226-12.783) |
|  | Multivariable | 1.001(1.000-1.002) | 1(reference) | 1.795(1.298-2.483) | 2.742(1.833-4.104) | 3.140(1.844-5.348) |

Odds ratios (95% confidence intervals) by multivariable logistic regression models were adjusted for age per year, sex, weight, RACHS-1, LVEF, baseline albumin, baseline serum creatinine, baseline hemoglobin.
RACHS, Risk Adjustment for Congenital Heart Surgery; LVEF, left ventricular ejection fraction.

## Data Availability

The data underlying this article and statistical analysis plan will be shared with researchers on reasonable request to the corresponding author. Participant data with identifiers are not openly available for access. Deidentified participant data supporting the findings of this study will be available at the time of publication. Please contact the Dr. Wang's for data-related queries or requests.

## Glossary of Abbreviations

AKI: acute kidney injury
KDIGO: Kidney Disease Improving Global Outcomes
OR: odds ratio
CI: confidence interval
SCr: serum creatine
CSA-AKI: cardiac surgery associated acute kidney injury
LVEF: left ventricular ejection fraction
RACHS-1: Risk Adjustment for Congenital Heart Surgery-1
ROC: receiver operating characteristics curve
CPB: cardiopulmonary bypass
HR: hazard ratios
ECMO: extracorporeal membrane oxygenation
RRT: renal replacement therapy

